# BCG protects against COVID-19? A word of caution

**DOI:** 10.1101/2020.04.09.20056903

**Authors:** Reka Szigeti, Domos Kellermayer, Richard Kellermayer

## Abstract

The COVID-19 pandemic, caused by type 2 Severe Acute Respiratory Syndrome Coronavirus (SARS-CoV-2), puts all of us to the test. Epidemiologic observations could critically aid the development of protective measures to combat this devastating viral outbreak. A recent publication, linked nation based universal Bacillus Calmette-Guerin (BCG) vaccination to potential protection against morbidity and mortality from SARS-CoV-2, and received much attention in public media, even before its peer review. We wished to validate the findings by examining the association between daily rates of COVID-19 case fatality (i.e. Death Per Case) /Days of the endemic [dpc/d]) and the presence of universal BCG vaccination before 1980, or the year of the establishment of universal vaccination. There was no significant association in either analysis. In this work we emphasize caution amidst the publication surge on COVID-19, and highlight the political/economical-, arbitrary selection-, and fear/anxiety related biases, which may obscure scientific rigor. It is underscored that physical (social) distancing (i.e. quarantine) and use of personal protective equipment (PPE) are the only epidemiologic measures (Iceland being a great example, where universal BCG vaccination policy was never in place), which consistently associate with successful counteraction of morbidity and mortality during the pandemic.

## Introduction

There is a current global crisis from the Coronavirus Disease of 2019 (COVID-19) pandemic https://www.cdc.gov/coronavirus/2019-ncov/about/index.html. COVID-19 is caused by type 2 Severe Acute Respiratory Syndrome Coronavirus (SARS-CoV-2), which is a medium-sized, enveloped, positive-stranded RNA virus of the *Coronaviridae* family. SARS-CoV-2 is the pathogen of the third, large, severe respiratory syndrome outbreak caused by Coronaviruses (CoVs) (1: SARS [severe acute respiratory syndrome] which emerged in late 2002 and disappeared by 2004; 2: MERS [Middle East respiratory syndrome], which emerged in 2012 and remains in circulation in camels] https://www.niaid.nih.gov/diseases-conditions/coronaviruses).

COVID-19 cases and associated deaths continue to exponentially rise https://www.worldometers.info/coronavirus/, which naturally induces fear, anxiety and sadness in all of us. Time is of essence towards finding definite solutions for stopping the pandemic, and scientists are under significant pressure trying to balance speed with safety and precision https://www.nbcnews.com/science/science-news/scientists-under-pressure-try-balance-speed-safety-coronavirus-vaccine-research-n1168946. Since fear and sadness can alter our cognitive control [1], there is valid concern about loosened scientific rigor in respect to the massive surge of publications amidst the time pressure on biomedical scientists racing for a cure. Even though the outbreak likely began in December of 2019 in Wuhan of Hubei Province in China [2], there are ongoing uncertainties about SARS-CoV-2 epidemiology. Recent rigorous studies (including multiple site and repeated nucleic acid based-, and also viral culture based testing) in 9 symptomatic patients with mild disease course have shown active viral replication in the upper airway, and high viral shedding in pharynx (but lower than in sputum) peaking at 4 days of symptoms [3]. SARS-CoV-2 virus was readily isolated from throat-and lung-derived samples, but not from stool, in spite of high virus RNA concentration in the fecal samples [3]. Blood and urine never yielded live virus [3]. These findings underscore the primary respiratory spread of

SARS-CoV-2, at least during mild disease course, which has been traditionally considered to be large droplet/contact communicated based on findings with SARS and MERS. However, some yet unpublished work indicates the potential airborne spread of the virus [4], leading to debates amongst infectious disease experts between contact vs. airborne protection recommendations. Adding to the difficulties in making clear cut regulations for personal protective equipment (PPE) use is viral shedding before symptomatic presentation [5]. Importantly, a large proportion of infected people can be asymptomatic as indicated from Iceland https://www.cnn.com/2020/04/01/europe/iceland-testing-coronavirus-intl/index.html, where the largest number of SARS-CoV-2 testing per million people has been performed to date. Such asymptomatic viral spreading may be most prominent in children, with over 50% being asymptomatic or having mild disease [6, 7]. Additionally, some patients present with gastrointestinal complaints, for example, and never develop respiratory symptoms https://journals.lww.com/ajg/Documents/COVID19_Han_et_al_AJG_Preproof.pdf. Furthermore, due to limitations in nucleic acid based analysis including quality of sample collection and variable methodology [8], a single nasopharyngeal swab can have as low as 32% sensitivity over the course of infection [9]. In the meantime, an asymptomatic patient may have similar viral loads as symptomatic ones, indicating the transmission potential from such cases [10]. These observations add to the tremendous difficulties in developing clear guidelines for PPE use during the COVID-19 pandemic, especially in the medical setting.

As already mentioned, similarly to MERS [11] and SARS [12], pediatric patients with COVID-19 run a much milder disease course than the elderly (especially above 60 years of age) [13]. The exact reason for this is unknown [7], but at least in non-human primates with experimental SARS-CoV infection, immune responses (most prominently CD8 T cell and B cell associated) were greatly reduced in the aged host compared to younger animals [14].

Consequently, a number of immunomodulatory treatments are being explored to help patients in fighting the infection. Amidst these explorations, prophylactic Bacillus Calmette-Guerin (BCG) vaccination in healthcare workers as a potential protection through immunomodulation against COVID-19 is being investigated in Australia’s Murdoch Children’s Research Institute https://about.unimelb.edu.au/newsroom/news/2020/march/preventative-vaccine-trial-for-covid-19-healthcare-workers. Linking to this study is a recently uploaded, not yet peer-reviewed paper [15] suggesting a connection between universal BCG vaccination policy and the peculiarly significant nation based variation in case frequency and death rates from COVID-19, based on data from March 21st, 2020. The epidemiology of the pandemic, however, is in ongoing flux. Therefore, we decided to re-examine this question with a modified definition of death rate based on April 3^rd^, 2020 data.

## Methods

COVID-19 epidemiologic data was extracted from https://www.worldometers.info/coronavirus/ on the afternoon (2 P.M. CDT USA) of April 3^rd^, 2020 for the top 68 countries based on number of cases. Day of country dependent “onset” was defined as first confirmed case reported and extracted from https://ourworldindata.org/coronavirus. Total number of days from day of onset to April 3^rd^, 2020 was calculated for each country. Due to tremendous variation in population based testing (36/million in Indonesia to 74,416/ million in Iceland) and the importance of time between diagnosis and death, we arbitrarily defined death rate as Death Per Case (i.e. case fatality)/Days (dpc/d) for the endemic of each country.

Data on BCG vaccination was extracted from the BCG World Atlas [16] similarly as in [15], or from online searches for those few countries, which were not analyzed in the Atlas, one example being Iceland [17]. Modern “Colonial Era” countries to colonize America and Africa were defined as: Netherlands, Spain, United Kingdom, France, Belgium, Portugal, and Germany.

Non-parametric Mann-Whitney U test was performed in Prism, Pearson correlation was calculated at https://www.socscistatistics.com/tests/spearman/default2.aspx. Graphs were created with Prism. Statistical significance was arbitrarily defined as p<0.05.

## Results

As opposed to Miller, et al. [15], we did not exclude countries with a population less than 1 million from our analyses, arguing that smaller countries may actually have better policies for universal testing (i.e. supporting rigorous epidemiologic analyses) than larger ones, Iceland being the prime example. Rather, we decided to study the top 68 countries for number of cases reported. This subjective cut-off was made for the Diamond Princess Cruise ship included in the list ranking at 68^th^ (the ship’s data was excluded). Amongst these 68 countries, we could identify the initiation year of universal BCG vaccination in 40 (Supplementary Table 1). Out of the countries examined, 9 did not have universal BCG vaccination before 1980 (Supplementary Table 1), which date we arbitrarily selected as the cutoff for having BCG vaccination “introduced” in respect to COVID-19 (since that would have affected the population of a country 40 years old and above [i.e. the population with increased vulnerability towards the infection]). There were no countries in our list, which had universal BCG vaccination in place and decided to stop that before 1980.

We first analyzed death rate according to Miller, et al. [15], namely death/million of the population. Similarly as they have found, death/million death rate was significantly higher in countries without universal BCG vaccination in place before 1980, compared to those which had (Figure 1A, p=0.001). Yet, there was no correlation (not shown, r_s_= −0.21632, p=0.18) between the year of the establishment of universal BCG vaccination and the mortality rate by death/million as opposed to the findings of Miller, et al. In any case, it is our strong impression that political and economic variations between countries significantly influence COVID-19 testing rates (including in those who are pre-terminal/critically ill). The lack of a single documented case from North Korea may represent such bias https://www.latimes.com/world-nation/story/2020-04-04/north-koreas-official-coronavirus-count-zero-why-that-claim-is-hard-to-believe. Additionally, the days into the country based epidemic from the first documented case can also significantly influence death rates. This is especially true in countries at the exponential phase of the outbreak, since critical condition usually develops 1-2 weeks after the onset of symptoms [18] (and presumptively after the diagnostic testing). Therefore, we defined death rate as Death Per Case (or case fatality rate)/Days from onset (dpc/d) to account for testing and time bias.

**Figure 1.**
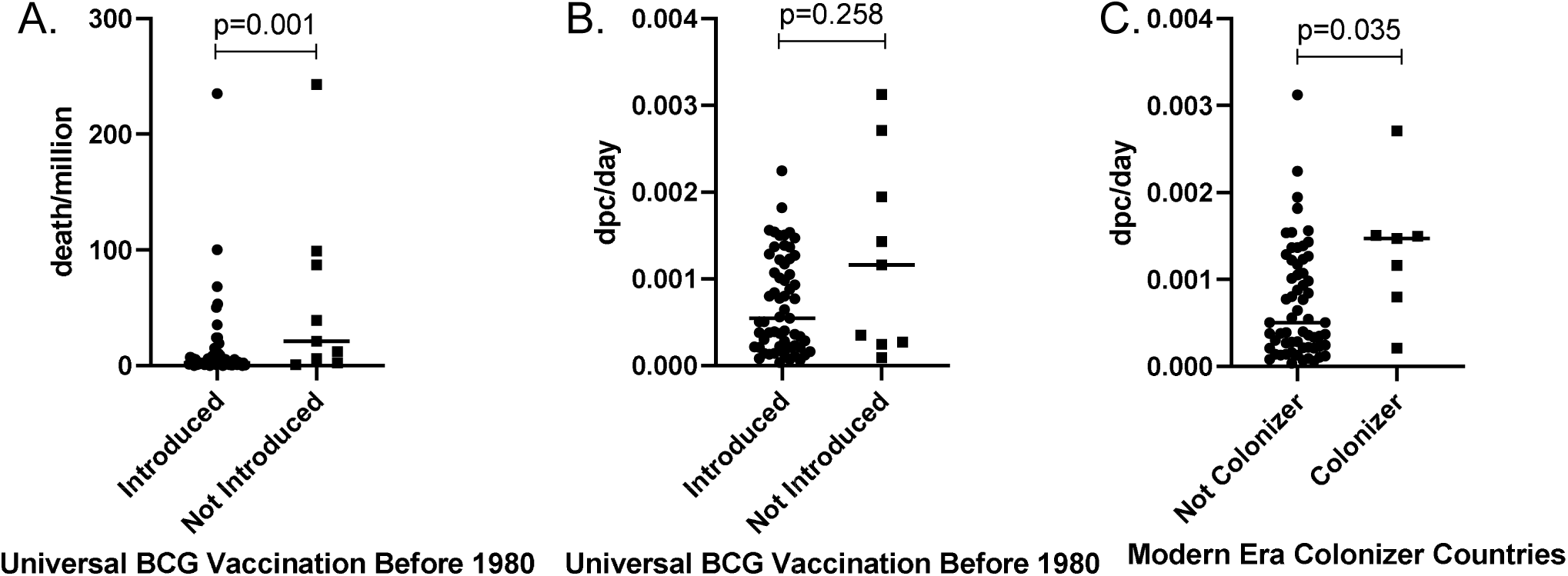
Association between COVID-19 mortality and country based variables. **A**. Death per million people in a country significantly associated with universal BCG vaccination present before 1980 (n=9) versus not (n=59). **B**. Death Per Case/Days of endemic (dpc/d), however, did not. **C**. Modern Era Colonizer countries (n=7) had higher death rates by dpc/d compared to those countries which are not (n=61). P values calculated by Mann-Whitney U test in Prism

Death rate according to dpc/d was not different between countries without universal BCG vaccination in place before 1980, compared to those which had (Figure 1B, p=0.258). Similarly there was no correlation (not shown, rs= −0.03136, p= 0.852) between the year of the establishment of universal BCG vaccination and the mortality rate by dpc/d. Furthermore, to underscore the potential for selection bias in this setting, we decided to examine dpc/d based death rates between countries partaking in the modern era colonization of America and Africa, and those which did not. Death rates were significantly higher in Modern Era Colonizer countries compared to those which were not (Figure 1C, p=0.035).

## Discussion

In this study, we found no association between universal BCG vaccination and country based COVID-19 mortality variation as defined by an arguably more precise death rate (i.e. dpc/d) definition than by Miller, et al. [15]. We show testing (political/economical), time (fear/anxiety), and selection (i.e. arbitrary selection giving rise to significant findings, such as Modern Era Colonizer countries, in our example) associated bias in such epidemiologic examinations. Speculative biologic explanation for Colonizer countries having higher mortality rates from COVID-19 could be generated (long standing, transgenerational influence of improved prenatal nutrition [19] on postnatal immune responses and life expectancy [leading to increased vulnerability to COVID-19] in these richer countries compared to the rest of the world, for example). Therefore, our work highlights the difficulties in drawing reliable epidemiologic conclusions from the currently available worldwide data on the COVID-19 pandemic. This conclusion is in line with experts in the field https://www.bbc.com/future/article/20200401-coronavirus-why-death-and-mortality-rates-differ, emphasizing that “the data is not from peer-reviewed research, but rather is almost real-time clinical data – which can be messy and come with many caveats”. The experts also underscore that “the lack of widespread, systematic testing in most countries is the main source of discrepancies in death rates internationally”. Indeed, it is the widespread testing and drastic quarantine measures for travelers that has led to exemplary flattening of the COVID-19 outbreak curve in Iceland https://www.icelandreview.com/ask-ir/whats-the-status-of-covid-19-in-iceland/, where there has never been a universal BCG testing policy in place. Additionally, the use of appropriate protective gear or personal protective equipment (PPE), especially in the healthcare setting has added to a similar success in South Korea and China https://www.thelancet.com/journals/lanres/article/PIIS2213-2600(20)30134-X/fulltext. The lack of strict home stay and distancing policy, however, has associated with a very recent surge of new COVID-19 cases in Japan https://abcnews.go.com/Health/coronavirus-live-updates-us-forces-japan-declares-public/story?id=69992372, in spite of the country having a universal BCG vaccination policy in place since 1942 (Supplementary Table 1). This surge follows a relatively prolonged flat curve of the outbreak since January 15^th^, 2020, which was likely the result of the culturally present interindividual physical distancing and already common facemask use in the large Japanese cities https://theprint.in/health/social-distancing-is-the-norm-in-japan-thats-why-covid-19-spread-is-slow-there/384498/. Japan is actually one of the most vulnerable countries to COVID-19 from other epidemiologic perspectives, such as the highest percentage of elderly population.

As far as the utility of strict PPE use amidst the pandemic, we highlight a video from an Italian hospital where no medical staff has been infected: https://www.youtube.com/watch?v=RsJJtI9bwwo

In conclusion, we advocate for the following based on our findings and review:

1. Widespread, national level testing for COVID-19 incorporating asymptomatic people, due to a large proportion of asymptomatic cases, especially in children. Based on such testing in Iceland, it is estimated that 1% of the population is, or has been infected around 33 days into the epidemic https://www.bbc.com/future/article/20200401-coronavirus-why-death-and-mortality-rates-differ. Repeated testing or the use of 2 different methodologies would be warranted [3], since a single nasopharyngeal swab can have low sensitivity [9]. The testing should ideally include immunoglobulin based seroconversion examinations to better clarify the epidemiology of COVID-19, and allow for potential return to normal living for those seroconverted.
2. Strict physical distancing supported by widespread testing results (the lack of such in Japan [less than 400 tests per million] associating with late surge of cases is an outstanding example supporting this conclusion).
3. Testing should be most widespread (i.e. both patients and all personnel physically present in an institution performing patient care) and physical distancing the most strict (which can only be achieved by variable degree, but universal PPE use) in the healthcare setting. This conclusion is based on observations that infection can be asymptomatic (i.e. patients with independent complaints can be infectious), or present with atypical symptoms (such as gastrointestinal complaints). Indeed, colleagues from New York City are emphasizing to “Assume everyone has the potential to harbor the virus, even when asymptomatic, and take the appropriate safeguards” https://journals.lww.com/ajg/Documents/COVID_NYC_AJG_Preproof.pdf. PPE policy should consider potential airborne spread of the virus [4] since there is no current evidence to contradict that.

## Data Availability

all data are publicly available

https://www.cdc.gov/coronavirus/2019-ncov/about/index.html

https://www.niaid.nih.gov/diseases-conditions/coronaviruses

https://www.worldometers.info/coronavirus/

https://www.nbcnews.com/science/science-news/scientists-under-pressure-try-balance-speed-safety-coronavirus-vaccine-research-n1168946

https://www.cnn.com/2020/04/01/europe/iceland-testing-coronavirus-intl/index.html

https://journals.lww.com/ajg/Documents/COVID19_Han_et_al_AJG_Preproof.pdf

https://about.unimelb.edu.au/newsroom/news/2020/march/preventative-vaccine-trial-for-covid-19-healthcare-workers

https://ourworldindata.org/coronavirus

https://www.socscistatistics.com/tests/spearman/default2.aspx

https://www.latimes.com/world-nation/story/2020-04-04/north-koreas-official-coronavirus-count-zero-why-that-claim-is-hard-to-believe

## References

1. Melcher T, Obst K, Mann A, Paulus C, Gruber O. Antagonistic modulatory influences of negative affect on cognitive control: Reduced and enhanced interference resolution capability after the induction of fear and sadness. Acta Psychol (Amst). 2012;139(3):507–14. Epub 2012/03/01. doi: 10.1016/j.actpsy.2012.01.012. PubMed PMID: 22366726.

2. Zhou P, Yang XL, Wang XG, Hu B, Zhang L, Zhang W, et al. Apneumonia outbreak associated with a new coronavirus of probable bat origin. Nature. 2020;579(7798):270–3. Epub 2020/02/06. doi: 10.1038/s41586-020-2012-7. PubMed PMID: 32015507.

3. Wolfel R, Corman VM, Guggemos W, Seilmaier M, Zange S, Muller MA, et al. Virological assessment of hospitalized patients with COVID-2019. Nature. 2020. Epub 2020/04/03. doi: 10.1038/s41586-020-2196-x. PubMed PMID: 32235945.

4. Santarpia JL, Rivera DN, Herrera V, Morwitzer MJ, Creager H, Santarpia GW, et al. Transmission Potential of SARS-CoV-2 in Viral Shedding Observed at the University of Nebraska Medical Center. medRxiv. 2020:2020.03.23.20039446. doi: 10.1101/2020.03.23.20039446.

5. He X, Lau EH, Wu P, Deng X, Wang J, Hao X, et al. Temporal dynamics in viral shedding and transmissibility of COVID-19. medRxiv. 2020:2020.03.15.20036707. doi: 10.1101/2020.03.15.20036707.

6. Qiu H, Wu J, Hong L, Luo Y, Song Q, Chen D. Clinical and epidemiological features of 36 children with coronavirus disease 2019 (COVID-19) in Zhejiang, China: an observational cohort study. Lancet Infect Dis. 2020. Epub 2020/03/30. doi: 10.1016/S1473-3099(20)30198-5. PubMed PMID: 32220650.

7. Dong Y, Mo X, Hu Y, Qi X, Jiang F, Jiang Z, et al. Epidemiological Characteristics of 2143 Pediatric Patients With 2019 Coronavirus Disease in China. Pediatrics. 2020. Epub 2020/03/18. doi: 10.1542/peds.2020-0702. PubMed PMID: 32179660.

8. Tyler AD, Smith MI, Silverberg MS. Analyzing the human microbiome: a “how to” guide for physicians. Am J Gastroenterol. 2014;109(7):983–93. Epub 2014/04/23. doi: 10.1038/ajg.2014.73ajg201473 [pii]. PubMed PMID: 24751579.

9. Wang W, Xu Y, Gao R, Lu R, Han K, Wu G, et al. Detection of SARS-CoV-2 in Different Types of Clinical Specimens. JAMA. 2020. Epub 2020/03/12. doi: 10.1001/jama.2020.3786. PubMed PMID: 32159775; PubMed Central PMCID: PMCPMC7066521.

10. Zou L, Ruan F, Huang M, Liang L, Huang H, Hong Z, et al. SARS-CoV-2 Viral Load in Upper Respiratory Specimens of Infected Patients. N Engl J Med. 2020;382(12):1177–9. Epub 2020/02/20. doi: 10.1056/NEJMc2001737. PubMed PMID: 32074444.

11. Thabet F, Chehab M, Bafaqih H, Al Mohaimeed S. Middle East respiratory syndrome coronavirus in children. Saudi Med J. 2015;36(4):484–6. Epub 2015/04/02. doi: 10.15537/smj.2015.4.10243. PubMed PMID: 25828287; PubMed Central PMCID: PMCPMC4404484.

12. Li AM, Ng PC. Severe acute respiratory syndrome (SARS) in neonates and children. Arch Dis Child Fetal Neonatal Ed. 2005;90(6):F461–5. Epub 2005/10/26. doi: 10.1136/adc.2005.075309. PubMed PMID: 16244207; PubMed Central PMCID: PMCPMC1721969.

13. Verity R, Okell LC, Dorigatti I, Winskill P, Whittaker C, Imai N, et al. Estimates of the severity of coronavirus disease 2019: a model-based analysis. Lancet Infect Dis. 2020. Epub 2020/04/03. doi: 10.1016/S1473-3099(20)30243-7. PubMed PMID: 32240634.

14. Clay CC, Donart N, Fomukong N, Knight JB, Overheim K, Tipper J, et al. Severe acute respiratory syndrome-coronavirus infection in aged nonhuman primates is associated with modulated pulmonary and systemic immune responses. Immun Ageing. 2014;11(1):4. Epub 2014/03/20. doi: 10.1186/1742-4933-11-4. PubMed PMID: 24642138; PubMed Central PMCID: PMCPMC3999990.

15. Miller A, Reandelar MJ, Fasciglione K, Roumenova V, Li Y, Otazu GH. Correlation between universal BCG vaccination policy and reduced morbidity and mortality for COVID-19: an epidemiological study. medRxiv. 2020:2020.03.24.20042937. doi: 10.1101/2020.03.24.20042937.

16. Zwerling A, Behr MA, Verma A, Brewer TF, Menzies D, Pai M. The BCG World Atlas: a database of global BCG vaccination policies and practices. PLoS Med. 2011;8(3):e1001012. Epub 2011/03/30. doi: 10.1371/journal.pmed.1001012. PubMed PMID: 21445325; PubMed Central PMCID: PMCPMC3062527.

17. Sigurdsson S. [Tuberculosis in Iceland. 1976]. Laeknabladid. 2005;91(1):69–102. Epub 2005/09/13. PubMed PMID: 16155306.

18. Goh KJ, Choong MC, Cheong EH, Kalimuddin S, Duu Wen S, Phua GC, et al. Rapid Progression to Acute Respiratory Distress Syndrome: Review of Current Understanding of Critical Illness from COVID-19 Infection. Ann Acad Med Singapore. 2020;49(1):1–9. Epub 2020/03/23. PubMed PMID: 32200400.

19. Waterland RA, Kellermayer R, Laritsky E, Rayco-Solon P, Harris RA, Travisano M, et al. Season of conception in rural gambia affects DNA methylation at putative human metastable epialleles. PLoS Genet. 2010;6(12):e1001252. Epub 2011/01/05. doi: 10.1371/journal.pgen.1001252. PubMed PMID: 21203497; PubMed Central PMCID: PMC3009670.

